# Rapid semi-quantitative point of care diagnostic test for post vaccination antibody monitoring

**DOI:** 10.1101/2021.09.17.21263729

**Authors:** Maria E. Moeller, Frederik N. Engsig, Melanie Bade, Jeppe Fock, Pearlyn Pah, Anna Louise Soerensen, Didi Bang, Marco Donolato, Thomas Benfield

**Affiliations:** Department of Infectious Diseases, Amager Hvidovre Hospital, Hvidovre, Kettegaard Allé 30, 2650 Hvidovre, Denmark; BluSense Diagnostics ApS, Symbion Bioscience Park, Fruebjergvej 3, 2100 Copenhagen, Denmark; Department of Clinical Microbiology, Copenhagen University Hospital, Amager and Hvidovre, Kettegaard Allé 30,2650 Hvidovre, Copenhagen, Denmark; Institute of Clinical Medicine, University of Copenhagen, Copenhagen, Denmark

**Keywords:** SARS-CoV-2, immuno-magnetic agglutination assay, vaccination, rapid IgG-IgM-IgA combined test, point-of-care, S protein trimer, total antibodies, spike trimer, vaccination monitoring

## Abstract

**Introduction:** **Point-of-care (POC)** quantification of the antibody responses against the SARS-CoV-2 Spike protein can enable decentralized monitoring of immune responses after infection or vaccination. We evaluated a novel POC microfluidic cartridge-based device (ViroTrack Sero COVID-19 Total Ab) for quantitative detection of total antibodies against SARS-CoV-2 Spike trimeric spike protein and compared to standard laboratory chemiluminescence (CLIA) based tests.

**Methods:** Capillary- and venous blood samples were collected from 101 individuals employed at Copenhagen University Hospital, Denmark. Antibody responses were measured on capillary-, venous whole blood, plasma and diluted plasma samples directly on the POC instrument. POC results were available within seven minutes on the microfluidic cartridge reader. Plasma samples were analysed on the DiaSorin LIAISON® XL CLIA Analyzer using LIAISON® SARS-CoV-2 IgM, LIAISON® SARS-CoV-2 S1/S2 IgG and LIAISON® SARS-CoV-2 TrimericS IgG assays. The data from the CLIA platform was used as a reference.

**Results:** The Spearman rank’s correlation coefficient between ViroTrack Sero COVID-19 Total Ab and LIAISON® SARS-CoV-2 S1/S2 IgG and LIAISON® SARS-CoV-2 TrimericS IgG assays is found to be 0.86 and 0.90 respectively. ViroTrack Sero COVID-19 Total Ab furthermore showed high correlation (>0.86) among the different sample matrixes. The agreement for determination of samples >200 BAU/mL on POC and CLIA methods is estimated to be around 90%.

**Conclusion:** ViroTrack Sero Covid Total Ab is a very rapid and simple-to-use POC test with high sensitivity and high correlation of the numerical results expressed in BAU/mL when compared to a commercial CLIA assay.

## INTRODUCTION

During coronavirus disease 2019 (COVID-19) pandemic, Severe acute respiratory syndrome coronavirus 2 (SARS-CoV-2) serological testing has been shown to play an important role not only as diagnostic support tool, but also in understanding antibody responses mounted upon SARS-CoV-2 infection and vaccination [1–3].

The spike (S) glycoprotein of SARS-CoV-2 forms surface-exposed homotrimers that mediate viral entry into host cells. Spiked glycoprotein is therefore the main target of SARS-CoV-2-specific neutralizing antibodies upon infection, and the focus of therapeutic and vaccine designs [4–6]. The correlates of protection are based on the specific level of SARS-CoV-2-specific neutralizing antibodies, acquired through vaccination or natural infection, that substantially reduces the risk of (re)infection [7,8].

Antibodies against the S protein are capable of neutralizing the virus and the S protein is therefore the primary antigen target of most of the current SARS-CoV-2 vaccines [9,10]. In clinical trials, antibody production and cellular T cell responses have been measured for these candidate vaccines [11–15]. It has been shown that a large proportion of the individuals undergoing seroconversion of immunoglobulin G (IgG) antibody responses against the viral S protein generate detectable neutralizing antibody responses [7], and that S protein binding assays correlate significantly with neutralization of wild-type SARS-CoV-2 virus [16–22]. Among the different subunits, the S protein in its trimeric form, when used in serology assays has a high sensitivity[23] and specificity[22].

Quantification of antibody responses and conversion rates of vaccinated populations can provide useful information not only to estimate the variety of vaccine responses and duration of protection, but also to enhance vaccine immunogenicity, dosage optimization, amount and time intervals [9,24]. Therefore, it is inevitable that SARS-CoV-2 S-based assays play an essential role in vaccine efficacy monitoring.

Several quantitative IgG or total antibody tests based on enzyme-linked immunoassay (ELISA) or chemiluminescence based instruments (CLIA) have been commercialized and their performances evaluated in depth [25–27]. However, none of these methods are applicable for antibody quantification in decentralized settings. Standardization of the First WHO International Standard for anti-SARS-CoV-2 immunoglobulin (human) (NIBSC code 20/136) has been introduced to allow for comparability between assay results. The International Standard is based on pooled human plasma from convalescent patients, which is lyophilized in ampules, with an assigned unit of 250 international units (IU) per ampule for neutralizing activity. For binding assays, a unit of 1000 binding antibody units (BAU) per ml can be used to assist the comparison of assays detecting the same class of immunoglobulins with the same specificity [28].

The threshold of protection for anti-SARS-CoV-2 S protein antibodies acquired by vaccination is an object of research in the recent phase of the pandemic. Initial studies show that antibody levels associated with immunity against symptomatic COVID-19 infection measures about 150-200 BAU /ml, using the WHO International Standard [8,29,30]. High antibody titers has been reported as above 250 BAU/ml [31]. Recent studies show correlations among antibody titers one month post-vaccination with the occurrence of breakthrough infections[32]; a third vaccine shot is the subject of discussion as it may boost immune systems and block new emerging coronavirus variants [33].

The aim of this study is to evaluate the performance of a new rapid quantitative point-of-care commercially available device from BluSense Diagnostics, based on the SARS-CoV-2 trimeric Spike protein, *ViroTrack® Sero COVID-19* Total Ab, with two CLIA laboratory-based immunoassays from Diasorin, *LIAISON® SARS-CoV-2 S1/S2 IgG* and *LIAISON® SARS-CoV-2 Trimeric S IgG* assay. The performance of the quantitative POC technology was evaluated on capillary- and venous blood drawn from 101 volunteers, employees from Hvidovre Hospital, Denmark.

## MATERIALS AND METHODS

### Subjects and samples

One hundred and one subjects were included during a three-day study in June 2021 (Characteristics of the study subjects are shown in Table 1). All participants were staff at the Hvidovre Hospital, Denmark, and included individuals from most types of professions (E.g. Doctors, Nurses, Physiotherapists, Cleaning Staff etc). Each study subject was asked to fill in a questionnaire in RedCap on demographics and vaccination status, previous infection with SARS-CoV-2 etc (Table 1). The study was approved by the Regional Ethics Committee of the Capital Region of Denmark (record no. H-20046624). The study was further approved by the Regional Data Protection Center (record no. P-2020-358).

**Table 1.**
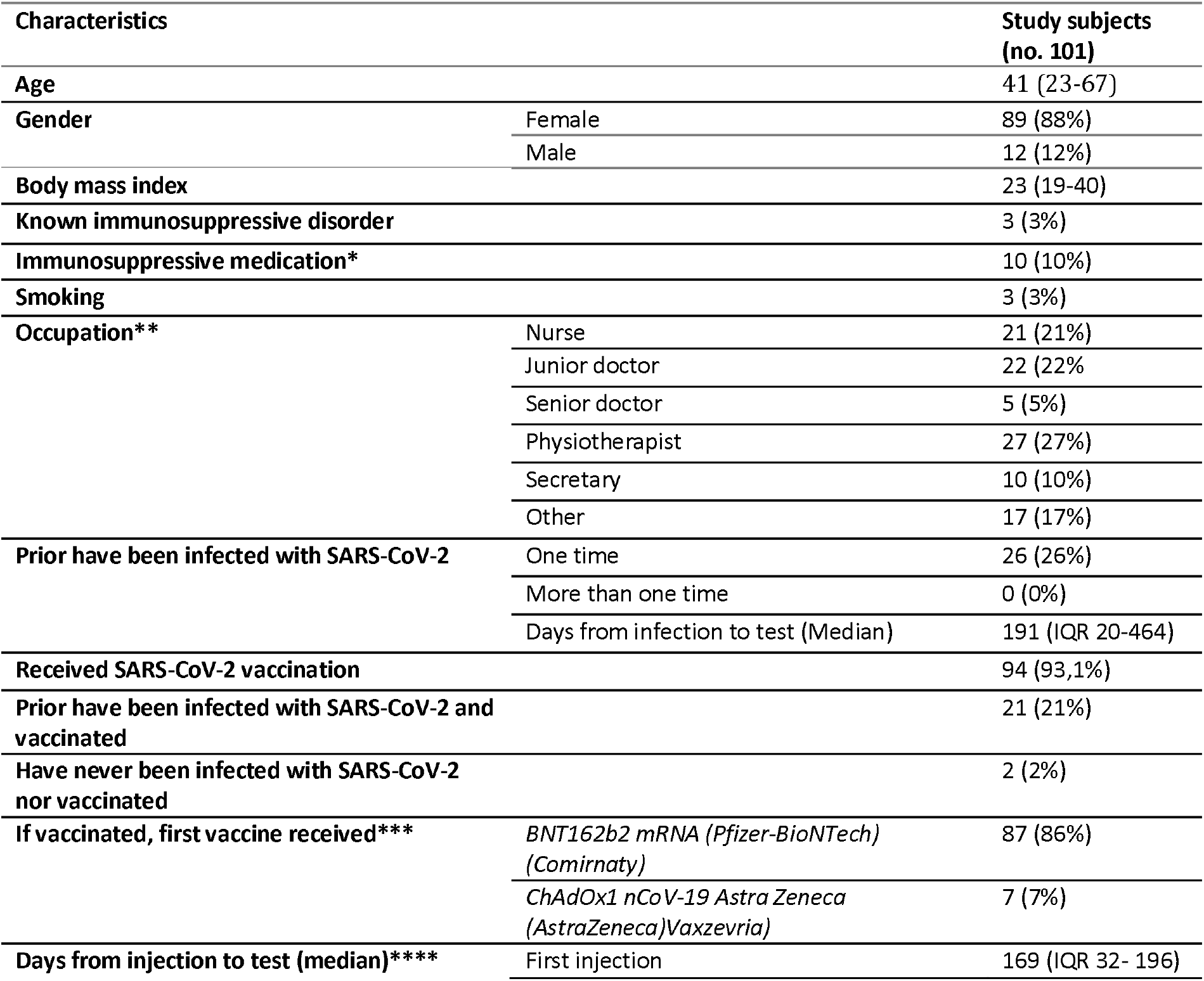
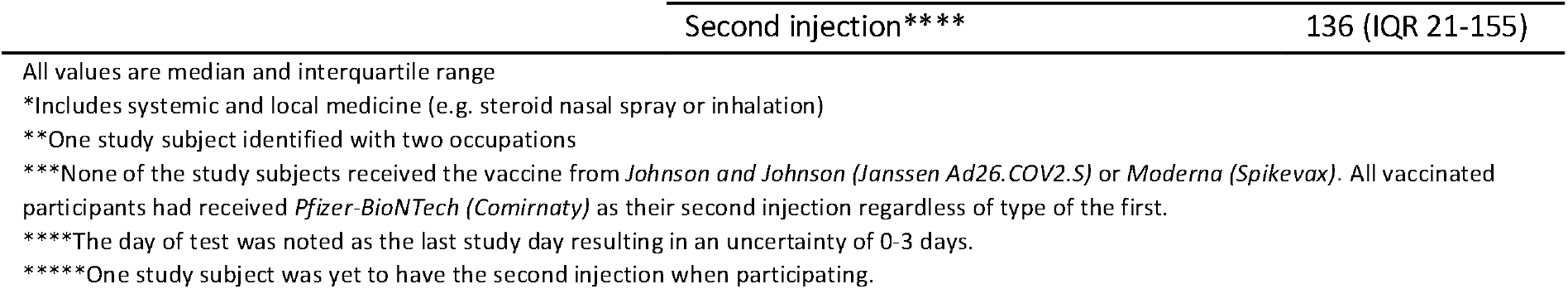
Characteristics of the study subjects

### Sample collection

Capillary- and venous blood sample were collected from each of the study subjects. Venous blood, capillary blood and plasma samples (diluted or undiluted) were analysed with the ViroTrack Sero COVID-19 Total Ab. The plasma samples were furthermore analysed on a Diasorin LIAISON® XL Analyzer at the Department of Clinical Microbiology, Hvidovre Hospital. Approximately 5 ml of venous blood was collected per subject in a blood collection tube (BD Vacutainer, UK) treated with potassium ethylenediaminetetraacetic acid (EDTA). Blood was processed following manufacturer’s instruction and used for plasma separation. Briefly, blood was centrifuged 15 minutes at 1500 x g and 20°C to obtain plasma.

For analysing capillary blood 20 µL was collected with a micropipette or a capillary pipette.

Capillary blood was loaded immediately onto the ViroTrack Sero COVID-19 Total Ab cartridge after collection. Venous blood was stored at room temperature after collection and to a maximal of 5 hours before testing on the ViroTrack Sero COVID-19 Total Ab cartridge and the plasma separation. Plasma was further stored at room temperature and to a maximal of 5 hours before testing onto the ViroTrack Sero COVID-19 Total Ab cartridge. All plasma samples were stored at -80 °Celsius prior to Diasorin LIAISON® XL Analyzer testing. For additional investigation and when necessary plasma was diluted in PBS at different dilution factors (1:10; 1:20; 1:40) and loaded onto the ViroTrack Sero COVID-19 Total Ab cartridge.

### ViroTrack Sero COVID-19 Total Ab

The ViroTrack Sero COVID-19 Total Ab is a POC rapid test providing quantitative results within seven minutes in the range 10-200 BAU/mL from 20 uL blood, plasma or serum. The test format is composed by a cartridge (ViroTrack Sero COVID-19 Total Ab) and a reader (BluBox).

The platform utilizes a centrifugal microfluidic platform together with an optomagnetic readout based on the agglutination of magnetic nanoparticles (IMA). In brief, 20 µl of sample is loaded on to the microfluidic cartridge which is then inserted inside the reader (BluBox). In the case of whole blood, the red blood cells are separated from the plasma by centrifugal force. The separated plasma is subsequently resuspended in the pre-stored reagents on the cartridge (e.g. magnetic particles). The magnetic particles are functionalized with SARS-CoV-2 trimeric spike protein and agglutinates in a sample containing anti-spike antibodies. Incubating the particles in a homogeneous magnetic field speeds up the reaction kinetics of the agglutination [34,35]. For optomagnetic detection, a uniaxial alternating magnetic field is applied which periodically aligns the agglutinated particle chains that results in a modulation of the transmitted light proportional with the target concentration [36]. IMA does not require labelled secondary antibodies [37].

The ViroTrack Sero COVID-19 Total Ab is calibrated to the to the “First WHO International Standard for anti-SARS-CoV-2 immunoglobulin (code: 20/136)” and the results converted in binding antibody units per milliliter (BAU/ml) by the software up to 200 BAU/mL. Plasma samples with >200 BAU/mL where diluted 10 times in PBS and re-measured. Dilution (20 times and 40 times) were continued until a result below 200 BAU/mL were obtained. The final binding antibody units per ml were found by multiplying the dilution factor with the obtained result. During the study four different BlueBox readers were used in parallel. Sample were loaded in different readers in randomized order.

### Diasorin

The plasma samples were analysed by chemiluminescence immunoassay (CLIA) for SARS-CoV-2 antibodies (IgM and IgG) targeting the subunits of the Spike proteins S1 and S2, the trimeric Spike complex or the receptor binding domain (S1-RBD). Samples were analysed on the DiaSorin LIAISON® XL Analyzer using LIAISON® SARS-CoV-2 IgM, LIAISON® SARS-CoV-2 S1/S2 IgG and LIAISON® SARS-CoV-2 TrimericS IgG assays. In accordance with a recent study on the specific assay, a negative SARS-CoV-2 result was defined as IgM index < 1.1 and IgG < 15 AU/mL, and a positive result was defined as an index value ≥ 1.1 AU/mL for IgM and ≥ 15 AU/ml for IgG, respectively. A TrimericS IgG result was defined as negative < 13 AU/ml, and a positive result defined as values ≥ 13 AU/ml (equivalent to ≥33.8 BAU/ml). The LIAISON® SARS-CoV-2 TrimericS IgG assay measures between 4.81 and 2080 BAU/mL. A recent study demonstrated that the DiaSorin SARS-CoV-2 S1/S2 IgG antibodies had a sensitivity of 96.2% and specificity of 98.9% [38]. Whereas, the DiaSorin TrimericS IgG has been shown to have a higher sensitivity of 99.4% and specificity of 99.8% [22].

### Statistical analysis

Spearman rank correlation was measured to evaluate the agreement between the different assays and the different specimen types. Analysis and graphs were performed using PYTHON /R.

## RESULTS

### Participant characteristics

A total of 101 participants were included. All characteristics can be found in the Table 1. Forty-seven-point-five percent of participants were between 20-39 years-old, 44.65% between 40-59 years-old and 8% were over 60 years of age. Of 101 participants, 93 were fully vaccinated (received two doses) and one had received only the first dose. Out of the fully vaccinated people, 52 participants received their second dose between two and up to five weeks after the first dose, and 41 participants five weeks (and up to 12 weeks) after the first dose. All the participants who received Astra Zeneca as the first dose, received BNT162b2 mRNA (Pfizer-BioNTech) as their second dose after at least 10 weeks. At the time of the study, all the participants who received BNT162b2 mRNA had received their second dose between 13 weeks to 29 weeks before the study and all the participants who received ChAdOx1 nCoV-19 (Astra Zeneca) had received their second dose between three to seven weeks before the study.

One capillary blood sample was not collected for one participant.

### Comparison of assay performances in plasma

Plasma was analysed with both the POC device and the central lab CLIA-based assays. All vaccinated individuals resulted in positive antibody titers with the ViroTrack system and both of the Diasorin IgG assays. Two participants whom were not vaccinated, but previously infected by COVID-19, were found to be negative by Diasorin TrimericS IgG, but to be low positive (20-50 BAU/mL) with ViroTrack for all specimen types.

Figure 1 shows the correlation between LIAISON® SARS-CoV-2 S1/S2 IgG, LIAISON® SARS-CoV-2 TrimericS IgG assays, LIAISON® SARS-CoV-2 IgM and ViroTrack Sero COVID-19 Total Ab for plasma samples. A strong correlation between ViroTrack and the Diasorin TrimiricS IgG and Diasorin S1/S2 IgG is observed. The Spearman rank’s correlation coefficient is found to be above 0.86 for the correlation between all three methods (see Table 1), and all the p-values are below <. The highest correlation with ViroTrack is obtained by the Diasorin Trimeric S assay. The Diasorin M assay do not display any correlation with the other tested methods.

**Figure 1:**
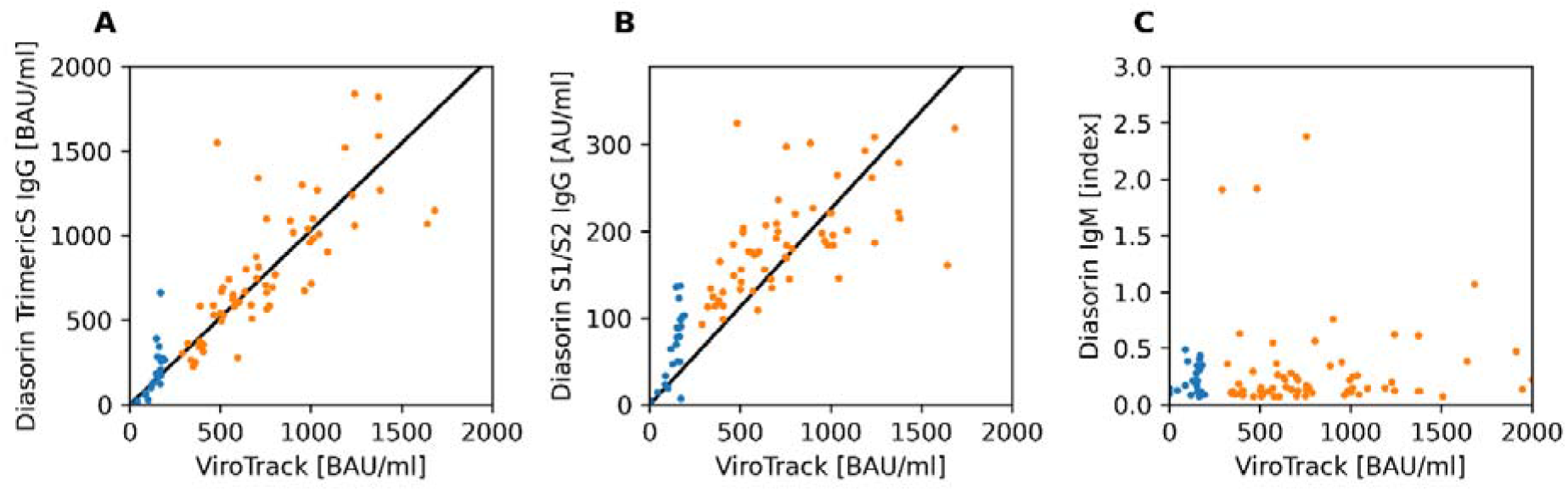
LIAISON® SARS-CoV-2 Trimeric IgG (A), S1/S2 IgG (B) and IgM (C) versus ViroTrack Sero COVID-19 Total Ab. For ViroTrack Sero COVID-19 Total Ab results are shown for diluted plasma (orange points) when >200 BAU/mL, otherwise undiluted results are shown (blue points). Results inside the dynamic range for Diasorin (<2080 BAU/mL for Trimeric IgG and <400 AU/mL for S1/S2 IgG) and below 2000 BAU/mL obtained for ViroTrack are shown.

**Table 1.** Spearman rank’s correlation coefficient. 16 samples above the dynamic range for Diasorin (2080 BAU/mL) are excluded from the analysis.

|  | ViroTrack | Diasorin TrimericS | Diasorin S1/S2 IgG | Diasorin IgM |
| --- | --- | --- | --- | --- |
| ViroTrack | 1.00 | 0.91 | 0.86 | 0.09 |
| Diasorin TrimericS | - | 1.00 | 0.93 | 0.14 |
| Diasorin S1/S2 IgG | - | - | 1.00 | 0.12 |
| Diasorin M | - | - | - | 1.00 |

### Comparison among capillary blood, venous blood and plasma results

Having established the correlation between ViroTrack and the reference test methods, the correlation between different specimen types is investigated. Figure 2 compare the results for different undiluted specimen types for values below 200 BAU/ml. The correlation between capillary blood, venous blood and blood plasma in the dynamic region (<200 BAU/ml) were high with Spearman rank’s correlation coefficients above 0.86 (Figure 2C).

**Figure 2.**
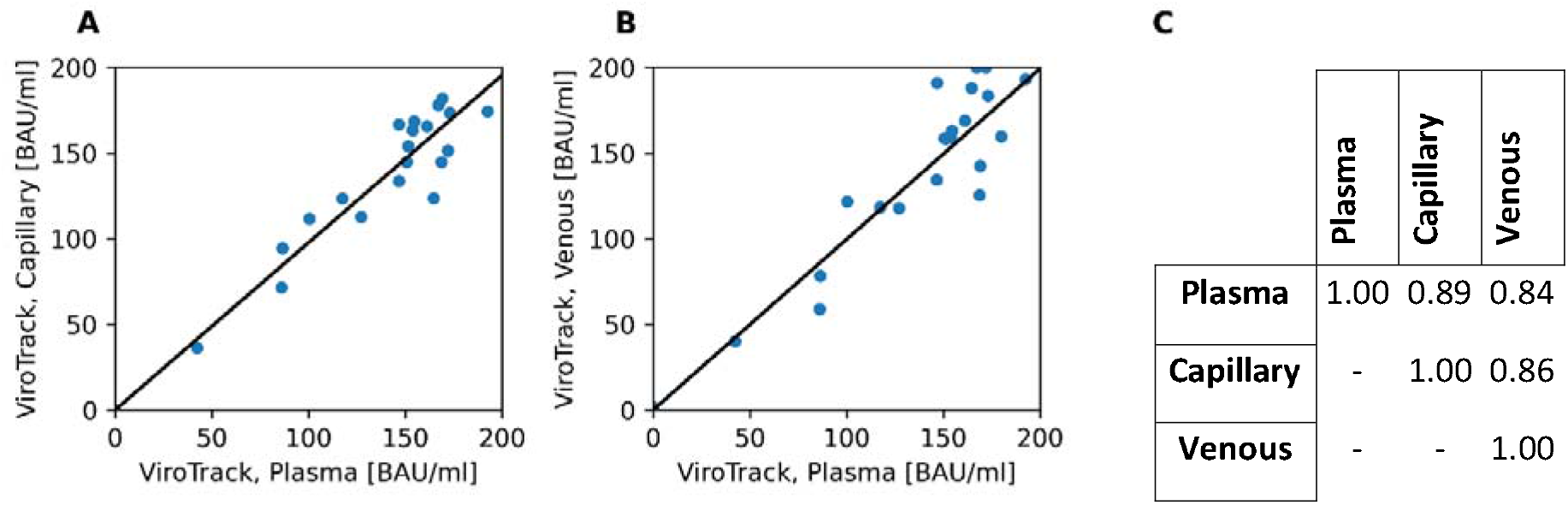
Specimen type agreement between ViroTrack Sero COVID-19 Total Ab results, using capillary, venous blood, and plasma samples. A) undiluted capillary blood versus undiluted plasma samples. B) undiluted venous blood versus undiluted plasma samples. C) Spearman rank’s correlation coefficient. Samples above 200 BAU/mL excluded from the analysis. All p-values are below

The capability of the assays to correctly quantify a high positive sample (e.g. >200 BAU/mL) can be important as revealed by recent reports correlating between the antibody levels and the immunity against symptomatic COVID-19 infection (Feng et al., 2021a; Khoury et al., 2021). The correlation between the different specimen types measured by ViroTrack and the plasma samples using Diasorin TrimericS IgG for determination of above 200 BAU/mL is demonstrated in table 3. The agreements are above 88% between all specimen types and both methods.

**Table 2.**
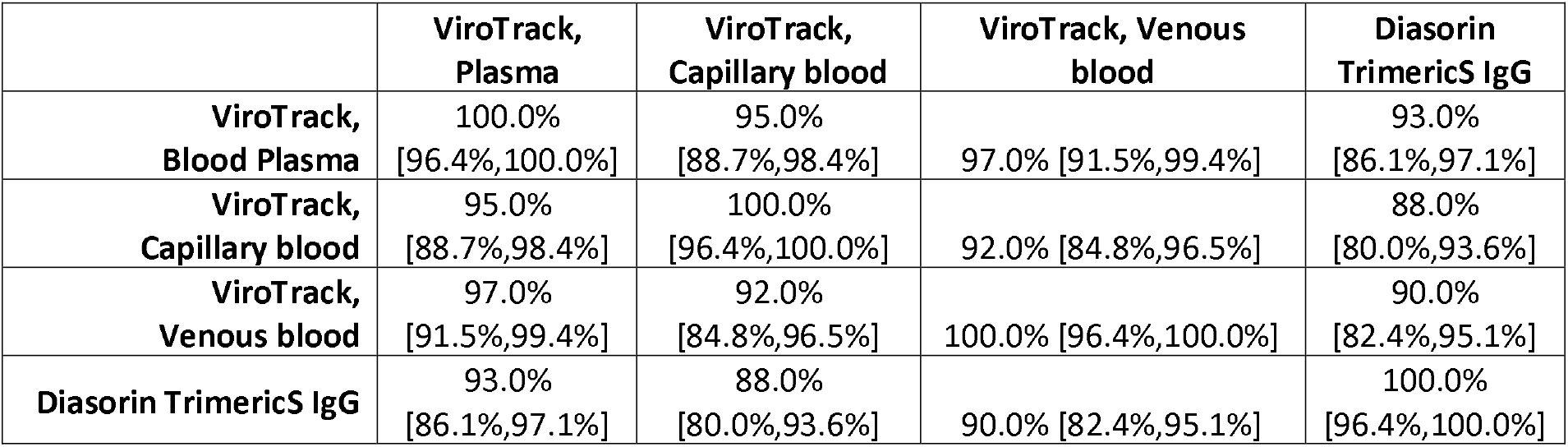
Agreement between specimen types and methods for determination of >200 BAU/mL in undiluted samples. Values in the square parenthesis are 95% confidence interval calculated using the Clopper-Pearson method.

|  | **ViroTrack,**<br>**Plasma** | **ViroTrack,**<br>**Capillary blood** | **ViroTrack, Venous**<br>**blood** | **Diasorin**<br>**TrimericS IgG** |
| --- | --- | --- | --- | --- |
| **ViroTrack,**<br>**Blood Plasma** | 100.0%<br>[96.4%,100.0%] | 95.0%<br>[88.7%,98.4%] | 97.0% [91.5%,99.4%] | 93.0%<br>[86.1%,97.1%] |
| **ViroTrack,**<br>**Capillary blood** | 95.0%<br>[88.7%,98.4%] | 100.0%<br>[96.4%,100.0%] | 92.0% [84.8%,96.5%] | 88.0%<br>[80.0%,93.6%] |
| **ViroTrack,**<br>**Venous blood** | 97.0%<br>[91.5%,99.4%] | 92.0%<br>[84.8%,96.5%] | 100.0% [96.4%,100.0%] | 90.0%<br>[82.4%,95.1%] |
| **Diasorin TrimericS IgG** | 93.0%<br>[86.1%,97.1%] | 88.0%<br>[80.0%,93.6%] | 90.0% [82.4%,95.1%] | 100.0%<br>[96.4%,100.0%] |

### Quantitative results vs vaccination/previous infectious status

The quantitative results obtained by the POC device has been analysed with respect to the vaccination and previous infected status of the participants. Figure 3 shows a swarm plot of the data. For the previous non-infected who were vaccinated we observe a lower antibody response (median: 569 BAU/ml) compared to the infected and vaccinated participants (median: 2000 BAU/ml), although for the previously infected we see a large spread of antibody responses (IQR 2651 BAU/ml compared to 566 BAU/ml). In general, we observe a higher antibody response for participants with ChAdOx1 nCoV-19 as the first dose and BNT162b2 mRNA as the second dose (median: 3988 BAU/ml) compared to participants receiving both doses with BNT162b2 mRNA (median: 638). However, these participants had all received the second dose with BNT162b2 mRNA less than 50 days before the study, whereas the participants receiving two doses of BNT162b2 mRNA in most cases received the second dose more than 125 days before the time of the study. The time between the injections also varied for the two groups.

**Figure 3.**
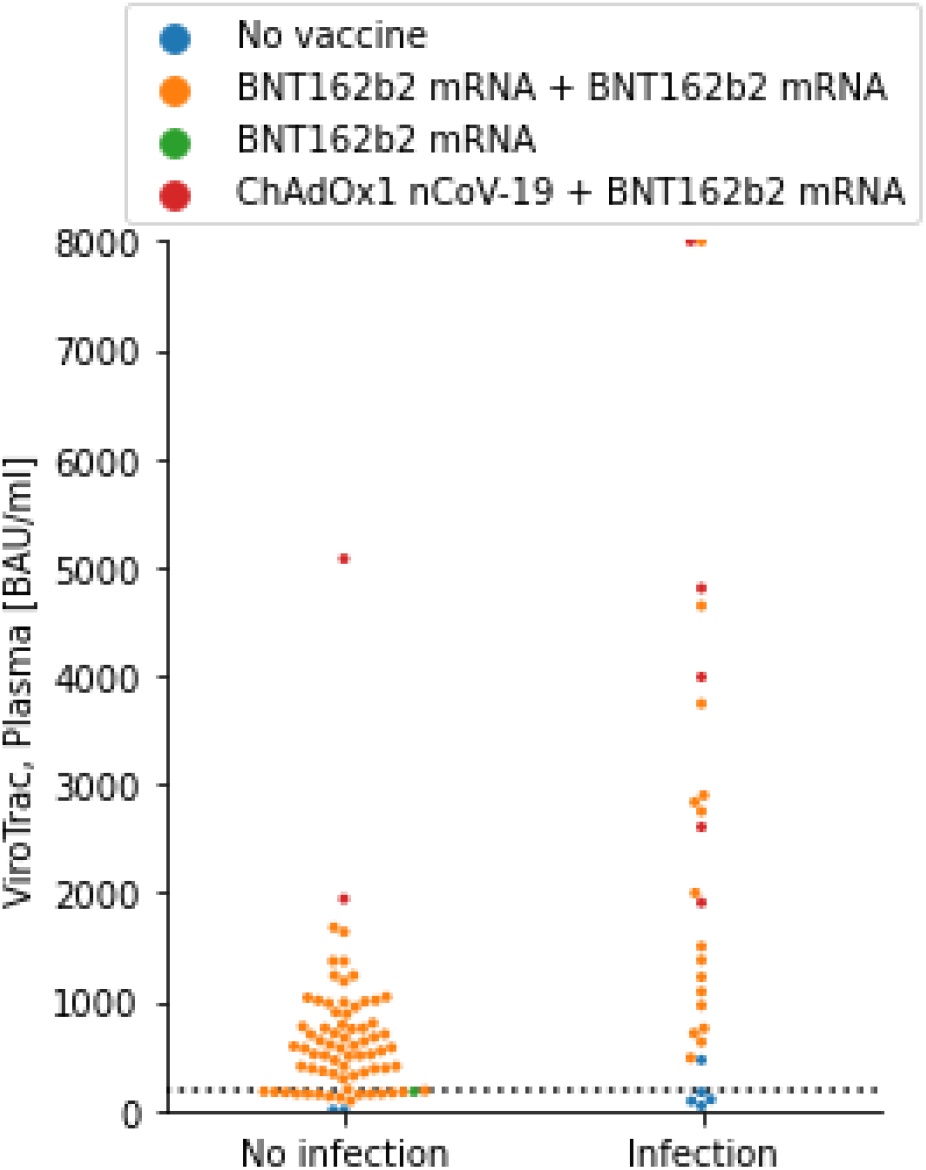
Swam plot of plasma and diluted plasma measured with ViroTrack Sero COVID-19 Total Ab. Data divided into previous PCR confirmed infected (Infection) and previous non-infected (No infection). The dotted line shows 200 BAU/mL.

Several of the non-infected who received both doses of BNT162b2 mRNA showed values below 200 BAU/mL. For the previous infected, only participants without any vaccination had values below 200 BAU/mL.

The study only contained two non-vaccinated and not previous infected participants (see **Error! Reference source not found**.). All methods and specimen types gave a negative test result for these two samples.

## DISCUSSION

To our knowledge, this is the first report comparing the numerical results of a rapid quantitative COVID-19 serology test with a reference CLIA method. Few semi-quantitative rapid serology tests generally based on fluorescence lateral flow tests with a reader exists, but so far only qualitative performances have been evaluated [39]. Our study showed a statistically high level of correlation (>0.9) between the results from *ViroTrack® Sero COVID-19* Total Ab and the two CLIA laboratory-based immunoassays from Diasorin, *LIAISON® SARS-CoV-2 S1/S2 IgG* and *LIAISON® SARS-CoV-2 Trimeric S IgG* assay. The highest correlation, 0.94, was been found with the *LIAISON® SARS-CoV-2 Trimeric S IgG* assay which utilises the same Spike trimer antigen as *ViroTrack® Sero COVID-19 Total Ab*.

Previous reports among various commercial Spike protein based IgG assays (ELISA or CLIA based) showed a lower level of correlation of around 0.7 to 0.8 [26] even when the results are translated in the BAU/mL units [25]. The high correlation is obtained even though the POC method measures total antibodies (IgG, IgM and IgA) while the reference methods measured only IgG and/or receptor binding domain. The low influence of IgA and IgM antibodies may be explained by a low IgM concentration, a general correlation between IgA and IgG titers or the predominance of IgG antibody class in vaccinated individual.

The agreement among specimen types was satisfactory. As described, the ViroTrack Sero COVID-19 Total Ab assay is embedded into a centrifugal microfluidics platform where blood is separated into plasma in the initial processing steps. This unique capability allows for the precise quantification without influence of factors as haematocrit, enabling precise correlation with laboratory-based methods. To our knowledge, systematic studies comparing COVID-19 antibodies in different matrixes does not exists, however preliminary studies shows differences in rapid test results when capillary or venous blood is used [40].

We observed a higher antibody response for participants with ChAdOx1 nCoV-19 as the first dose and BNT162b2 mRNA as the second dose less than 50 days prior to the study, compared to the participants who in most cases more than 125 days prior to the study received their second dose of their two doses from BNT162b2 mRNA [41,42].

A limitation of this study is represented by the fact that only two individuals were not infected nor vaccinated; it is therefore not possible to draw any conclusions on the specificity of the POC device vs the reference technique. In a previous study we showed that a first version of the POC test targeting the antibodies against SARS-COV-2 nucleocapsid protein to have higher specificity compared with ELISA based methods[37]. Secondly, the study is limited as an extra dilution step was necessary to extend the current dynamic range of the POC device. Which is currently not included in the product “instructions for use” and the dilution process performed in blood may have produced a different result. However, the data demonstrated that the device produce accurate quantification of diluted plasma.

A general agreement >90% between capillary blood, venous blood, and plasma from the same samples and techniques has been found thus supporting the use of capillary blood on the POC device for precise decentralized antibodies monitoring post-vaccination and responses after natural infection in countries where the use of vaccines is low or yet to come.

Among the vaccinated only individuals, 17 (18%) had antibodies below 200 BAU/ml.

Their median time from last vaccination to antibody test was 136 days (IQR; 138.5 -130.5) compared to 136 days (IQR; 139 – 130) in those who had above 200 BAU/ml.

The major limitation of our study is the lack of negative controls why we could not determinate specificity and sensitivity. Still, our result show high correlation with a commercial CLIA assay.

None of our assays included antibodies against other isotopes than the spike protein and information on previous infection with SARS-CoV-2 therefore relied on participant memory which could introduce a bias. On the other hand the main purpose of the study was to evaluate the performance of a new technology and our result should not be affected by this.

In conclusion, *ViroTrack® Sero COVID-19* Total Ab provides an accurate numerical quantification of the total antibodies against the Spike protein trimer within seven minutes from a single drop of capillary blood. Compared to rapid lateral flow tests detecting antibodies against different forms of the Spike protein the evaluated POC device provides a numerical result in a shorter time.

This capability can enable precise monitoring of antibodies amounts in facilities in various places allowing a potential wider use of quantitative serology tests in the COVID-19 pandemic.

## Data Availability

Data available upon request

## ACKNOWLEDGEMENTS

Heike Ebermann from the department of Microbiology, Copenhagen University Hospital, Amager and Hospital, for analyzing the samples on the Diasorin. Laura Catalina Bohorquez for the help in drafting the paper introduction.

